# St George’s COVID shield for use by ENT surgeons performing tracheostomies

**DOI:** 10.1101/2020.05.04.20087072

**Authors:** Sabrina Brar, Jahan Daya, James Schuster-Bruce, Sanjeev Krishna, Hamid Daya

**Affiliations:** ENT Department, St George’s University Hospital, London; Institute for Infection and Immunity, St George’s University of London

**Author notes:** **Acknowledgements** We would like to thank Matthew Buckley, Abplas, for his design skills, time and determination to help create the St George’s shield. We would also like to thank the ENT surgeons and theatre staff at St George’s University Hospital, for using and providing feedback on the shield. Our thanks also to Mr. Darren Lui, Trauma and Orthopaedic Surgeon, St George’s University Hospital, for sharing his experience of and allowing us to use his particle counter.

**Keywords:** COVID-19, coronavirus, SARS-CoV-2tracheostomy, ENT, aerosol-generating, PPE, St George’s shield

## Abstract

Healthcare workers are at increased risk of exposure to COVID-19. The majority of cases are acquired through inhalation of infected respiratory particles, contamination with infected surfaces or whilst performing aerosol-generating procedures. Basic infection prevention measures are essential to protect healthcare workers from contracting the disease when managing patients; consequently global demand for personal protective equipment (PPE) has exceeded supply in many regions. We present a novel, innovative polycarbonate shield designed for ENT clinicians performing tracheostomies. Clinical investigations using the shield demonstrated a sixteen-fold decrease in the number of particles detected at the position of the operating surgeon when the shield was used (particle size 0.3μm; with shield 27,000 versus 439,000 without shield). The shield, used with appropriate PPE, could therefore help to minimise exposure to aerosol generated particles such as during tracheostomies on patients with COVID-19.

## 1 Introduction

The COVID-19 pandemic began in late 2019 and has caused an estimated two hundred and twenty thousand deaths in over two hundred countries by the end of April 2020.^1^ This devastating spread is largely due to its relatively high transmissibility with an estimated mean (95% CI) R0 of 2.65 (1.97, 3.09).^2^ COVID-19 is primarily a pneumonic illness.

Airborne mechanisms of transmission vary depending on particle size. Larger particles (>5μm diameter) are called droplets.^3^ Droplet spread occurs when droplets containing coronavirus are expressed from an infected individual through a cough or sneeze to make contact with exposed mucosal surfaces, either directly, or from contact with contaminated surfaces. Aerosols or droplet nuclei are smaller particles (<5μm) that due to their smaller size are able to remain airborne, transmitting the virus if they are inhaled into the lungs.^3^ Aerosol transmission can occur up to 3 hours after the release of viral particles.^4^ Ninety-seven percent of SARS-CoV-2 aerosol particles have an aerodynamic diameter of less than 1μm (range 0.1μm to 900μm) and a dominance of between 0.25μm and 0.5μm.^5, 6^

Airway procedures that generate aerosols, such as tracheostomy, increase the risk of disease spread from infected patients. Tracheostomy is required for a significant proportion of ventilated COVID-19 patients weaning from ventilation. In studies of similar coronaviruses, tracheal intubation and tracheostomy were found to be associated with a 6-fold and 4-fold increased risk respectively, of transmission to healthcare workers.^7^

We hypothesise that barrier protection, with an aspiration unit, may decrease the number of particles that clinicians performing tracheostomies are exposed to, compared to no barrier at all. To test this hypothesis, we performed a clinical simulation on a hospital ward using a novel protective barrier, the St George’s shield. The St George’s shield is a bespoke barrier protection that encases the patient undergoing a tracheostomy.

## 2 Materials and Methods

### Ethical considerations

Consent was gained from our volunteer prior to them partaking in this clinical investigation. The volunteer used their own vaporising device (Vaporesso Swag II) for the purpose of the experiment.

### Study design

This was a cross sectional study design adhering to STROBE guidelines.^8^

### Setting

The study was performed on a standard hospital ward at St George’s University Hospital, London, in April 2020.

### Participants

One participant volunteered for the investigation. They were healthy, asymptomatic, and a non-smoker.

### Intervention

#### Method 1

The participant was positioned supine on a standard hospital bed in a hospital ward heated to 23.5°C. (Figure 1) They were asked to inhale one puff (equivalent to one deep breath) from the vaporiser; they then exhaled. The participant then simulated a cough to generate the release of aerosol particles, similar to when the trachea is opened during a tracheostomy. A particle counter detected the burden of particle generation. Particles were detected at different locations to represent the locations of different members of the operating team. (Figure 2a, 2b) The procedure was repeated 2 times with the St George’s shield and 2 times without.

**Figure 1.**
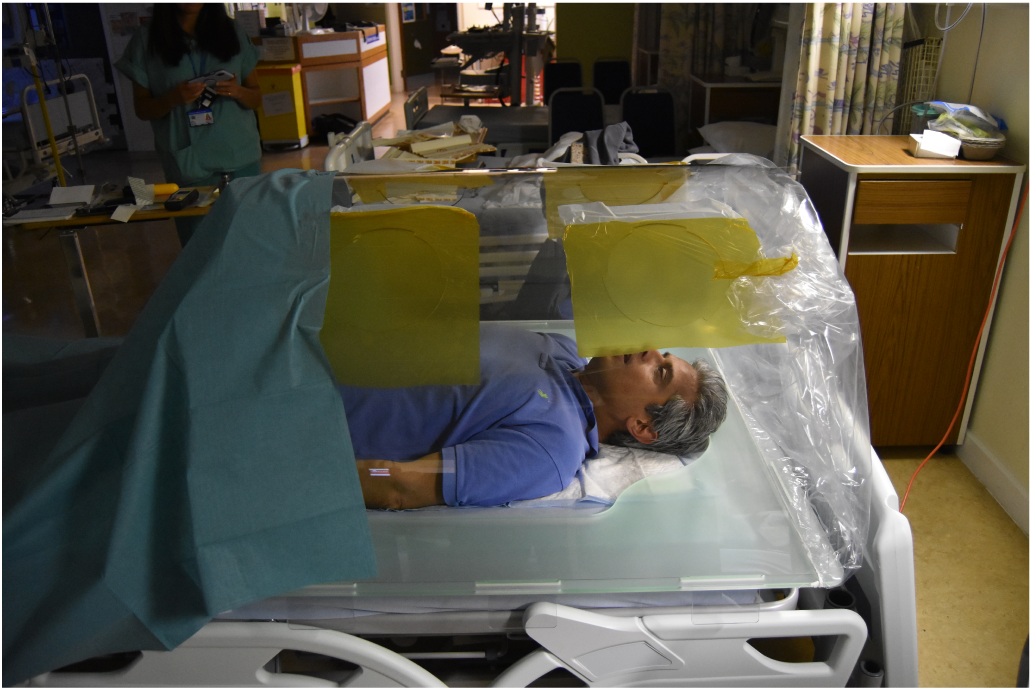
Simulation set up – the volunteer lies on the hospital bed with an assembled St George’s shield

**Figure 2a.**
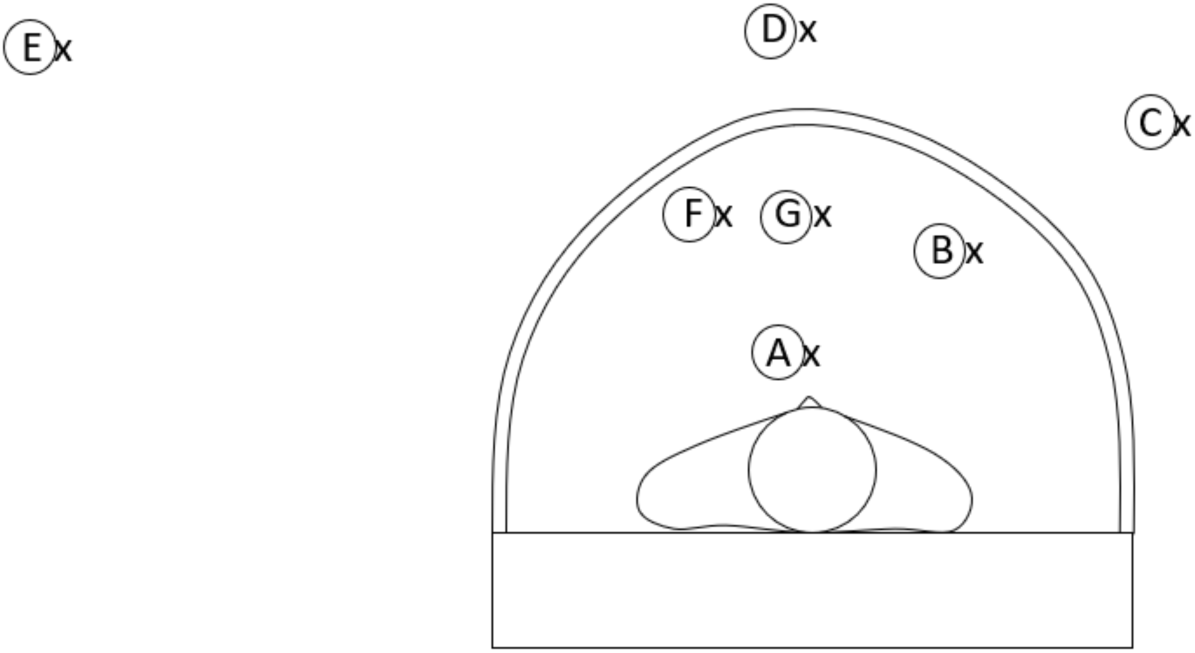
This shows the different locations of the particle detector. End-on view.

**Figure 2b.**
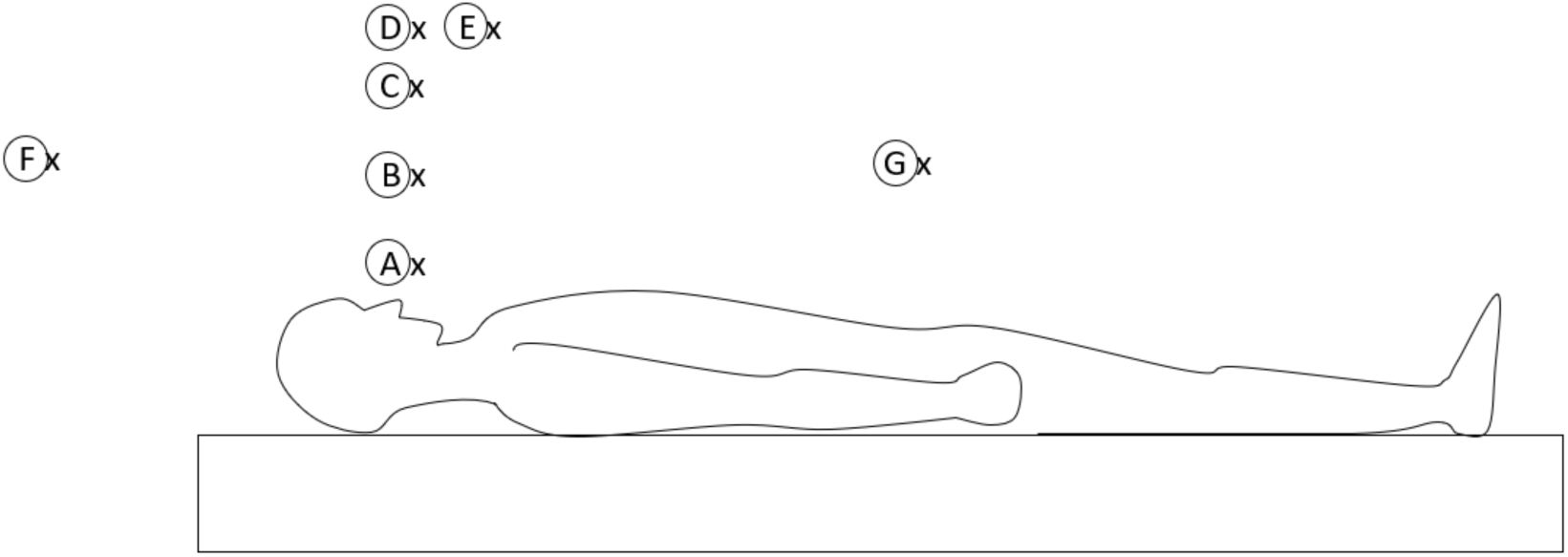
This shows the different locations of the particle detector. Side-on view.

#### Method 2

One further procedure was conducted whereby the volunteer was asked to inhale and exhale one puff from the vaporiser prior to *every* measurement of particles at different locations.

To analyse the risk associated with particles being contained within the shield, the coughing simulation was performed whilst particle measurements were taken at different time intervals over a period of 10 minutes. The effect of using an aspiration unit on particle measurements over time was also analysed, by placing a hoover hose within the shield.

Key for Figures 2b and 2b
A – Immediately above the volunteer’s mouth – represents the maximum number of particles released from coughing
B – Immediately under the shield’s surface (or equivalent without the shield) – measurement of the particles that are likely to adhere to the inside of the shield’s surface
C – The location of the operating surgeon’s head
D – Immediately above the shield – measurement of particles here demonstrated the efficacy of the shield
E – Two metres away from the volunteer, at standing head height – calculation of particles that might reach standing theatre staff
F – Anaesthetist
G – Immediately outside the shield at the foot end of the patient – this represents the location of the scrub nurse

The St Georges shield is a made up of 5 components. (Figure 3) 1) A polycarbonate base that sits around the patient with slots on either side. 2) A polycarbonate bendable sheet that acts as the shield, with ‘feet’ that slot into the openings in the base. 3) Domed side supports that provide stability to the main sheet. 4) Four iris port sleeves for the surgeon’s arms to fit through. 5) Four rings that connect these sleeves to the shield. In addition, two low fluid cloth drapes are attached to either end of the shield to complete the enclosure around the patient. Its ‘flat-pack’ design facilitates simple assembly, transportation and storage. The sterilisation regimen involves a ‘two wipe system’ whereby it is cleaned (PDI Sani Cloth detergent wipe - yellow top) and then sterilised (PDI Sani Cloth 70% alcohol wipe - red top). This was approved locally by the Infectious Diseases Department at our institution.

**Figure 3.**
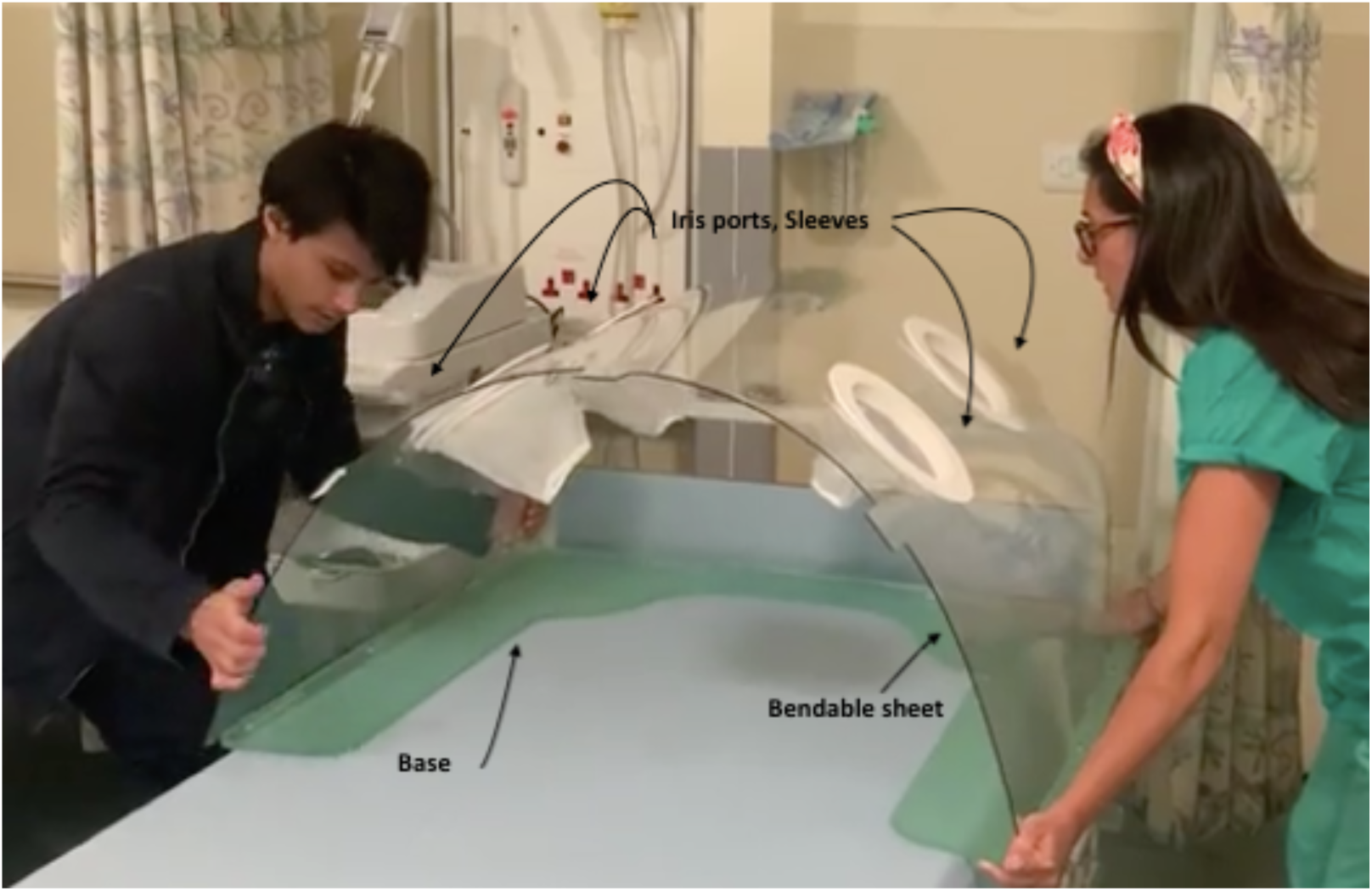
Assembly of the polycarbonate base, bendable sheet, iris ports and sleeves to form the shield. (The domed side supports are not seen in this image)

## Variables

The primary outcome was to compare the effect of the shield on particle frequency (according to particle size) at different locations during a simulated aerosol generating procedure.

The secondary outcome was to evaluate the length of time that particles remained within the device, both with and without an aspiration unit.

## Data sources / measurement

A particle counter (Air Quality Detector HT-9600/HT-9601 Particle Counter) was used to detect particle size and frequency. It was able to detect a range of three different particle sizes (0.3μm, 2.5μm, 10μm). The smallest sized particle, 0.3μm, is roughly equivalent to the size of SARS-CoV-2 aerosol particles.^5^ Analysis of the larger particle sizes illustrates the distribution of particles that may cause droplet transmission of COVID-19.

## Analysis

Data were analysed using Microsoft ^®^ Excel ^®^ for Mac 2011, Version 14.7.3.

## 3 Results

The average number of ambient particles measured prior to the start of the investigation was 27,000 for 0.3μm particles, 30 for 2.5μm and 0 for 10μm.

### Particle detection with and without the St George’s shield

The number of particles measured at positions A and B were equivocal with and without the shield for all three sizes of particles, using method 2. Locations C, D, E, F, G had notable differences in particle detection; average numbers of particles for method 1 ranged from 28.000 – 56,500 without the shield compared with 22,000 – 29,000 with the shield (for particles sized 0.3μm). (Table 1a, Figure 4) Difference in particle frequency was even greater using method 2 (particle frequency ranged from 22,000 – 1,245,000 without the shield versus 20,000 – 26,000 with the shield). (Figure 5) The greatest difference in particle detection using methods 1 and 2 was noted at position D (method 1 difference = 34,500 particles; 56,500 without the shield versus 22,000, with the shield; method 2 difference = 1.387.000 particles; 1,413,000 without the shield versus 26,000 with the shield). (Table 1a and 1b) Similar comparisons can be made with the larger particles.

**Figure 4.**
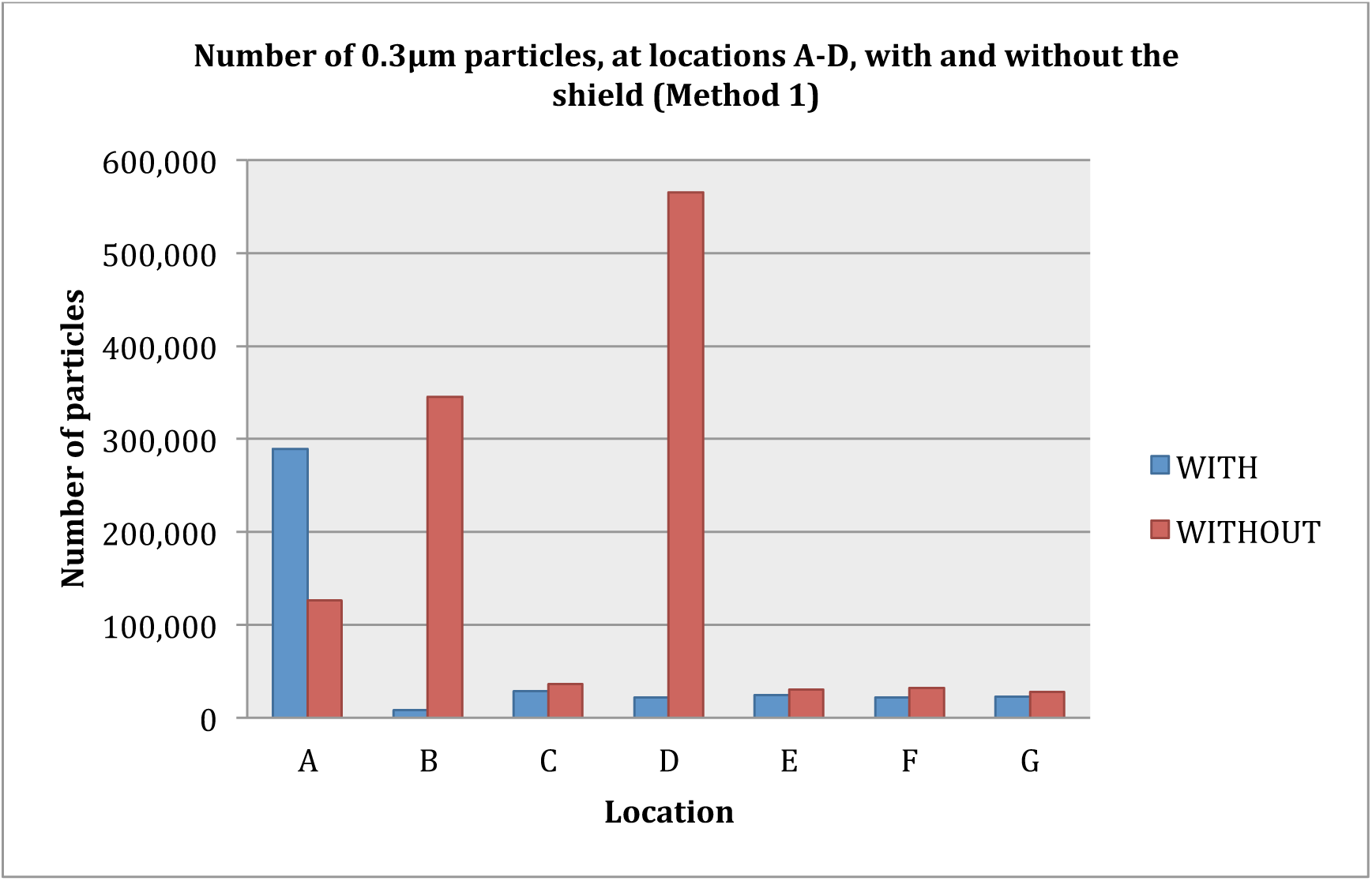
Graph showing the impact of the shield on detection of 0.3μm size particles using method 1 (average numbers)

**Figure 5.**
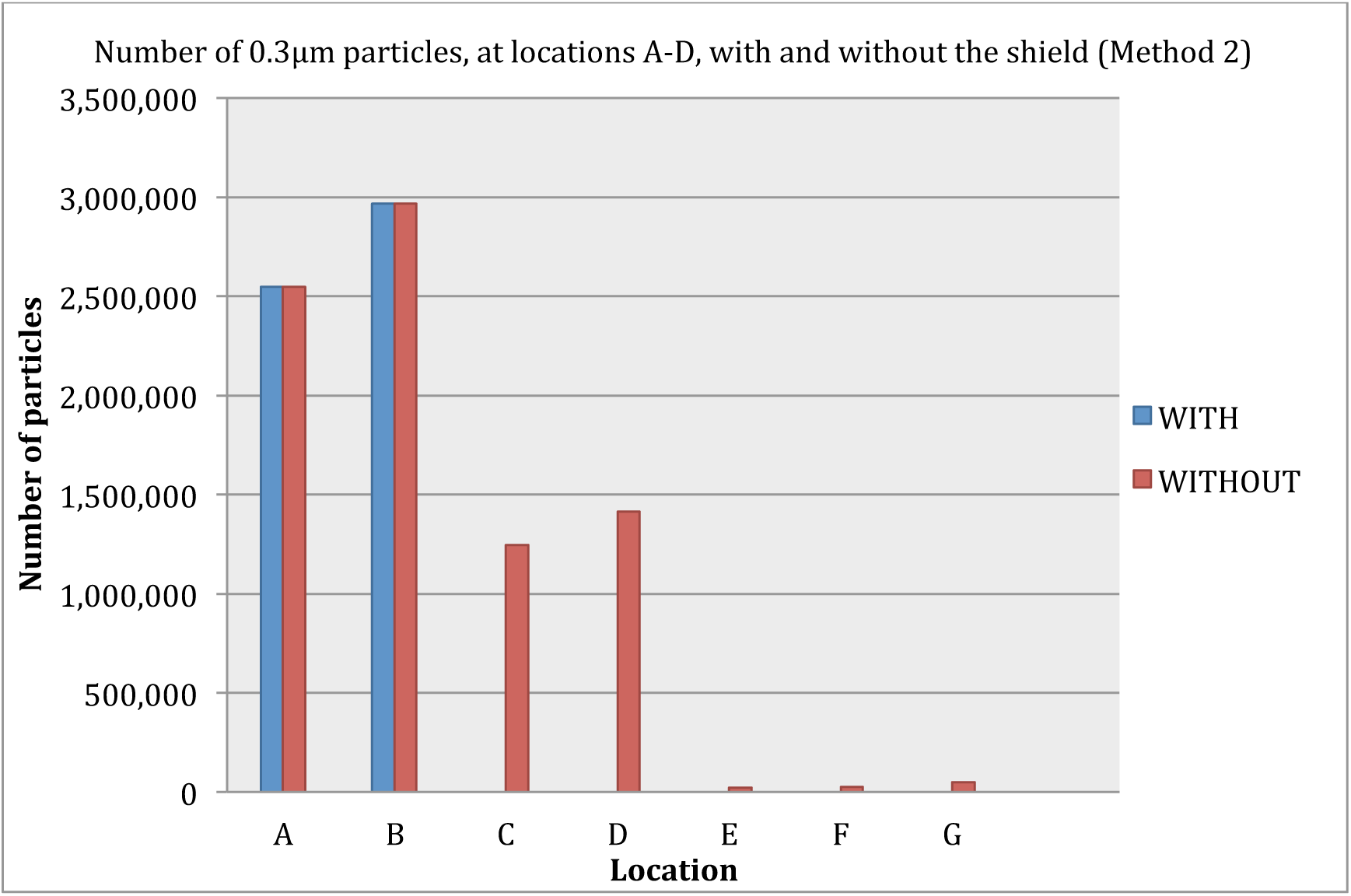
Graph showing the impact of the shield on detection of 0.3μm size particles using method 2

**Table 1a.** Average particle measurements taken at different locations, with and without the shield, using method 1

| Particle Size<br>(µm) | 0.3 |  | 2.5 |  | 10 |  |
| --- | --- | --- | --- | --- | --- | --- |
|  | With<br>shield | Without<br>shield | With<br>shield | Without<br>shield | With<br>shield | Without<br>shield |
| Location |  |  |  |  |  |  |
| A | 289,000 | 126,000 | 805 | 315 | 10 | 2 |
| B | 80,000 | 34,500 | 220 | 40 | 16 | 0 |
| C | 29,000 | 36,000 | 40 | 50 | 0.5 | 0 |
| D | 22,000 | 56,500 | 25 | 50 | 0 | 0 |
| E | 24,500 | 30,000 | 30 | 40 | 0 | 0 |
| F | 22,000 | 32,000 | 30 | 55 | 0 | 0 |
| G | 23,000 | 28,000 | 45 | 30 | 0 | 0 |

**Table 1b.** Particle measurements taken at different locations, with and without the shield, using method 2

| Particle Size<br>($\mu\text{m}$) | 0.3 | | 2.5 | | 10 | |
| --- | --- | --- | --- | --- | --- | --- |
|  | With<br>shield | Without<br>shield | With<br>shield | Without<br>shield | With<br>shield | Without<br>shield |
| Location |  |  |  |  |  |  |
| A | 2,549,000 | 2,549,000 | 15,860 | 15,860 | 828 | 828 |
| B | 2,967,000 | 2,967,000 | 17,870 | 17,870 | 990 | 990 |
| C | 23,000 | 1,245,000 | 40 | 3010 | 0 | 174 |
| D | 26,000 | 1,413,000 | 40 | 4000 | 0 | 224 |
| E | 20,000 | 22,000 | 20 | 60 | 0 | 0 |
| F | 25,000 | 27,000 | 20 | 30 | 0 | 0 |
| G | 21,000 | 48,000 | 40 | 2200 | 0 | 26 |

### Particle detection within the shield over time

Table 2 shows that over a 10-minute period, there was a reduction in the number of particles. At 10 minutes, the total number of particles (size 0.3μm) measured within the shield was less than measured at ambient levels (19,523 compared with 27,000). There were no particles sized 10μm detected within the shield after 10 minutes.

**Table 2.** The number of particles detected inside the shield at varying time intervals

| Time (minutes) | Particle size (µm) |  |  |
| --- | --- | --- | --- |
|  | 0.3 | 2.5 | 10 |
|  | Number of particles |  |  |
| 1 | 4700000 | 89000 | 2499 |
| 2 | 4758000 | 78000 | 2548 |
| 3 | 4520000 | 64800 | 2178 |
| 4 | 3166000 | 35000 | 1441 |
| 5 | 2730000 | 25000 | 997 |
| 10 | 19523 | 10 | 0 |

### Particle detection within the shield with an aspiration unit over time

The aspiration unit reduced the number of particles detected within the shield over time. 22,000 particles (ambient level) sized 0.3μm were measured under the shield after 1 minute when the aspiration unit was used. (Table 3)

**Table 3.**
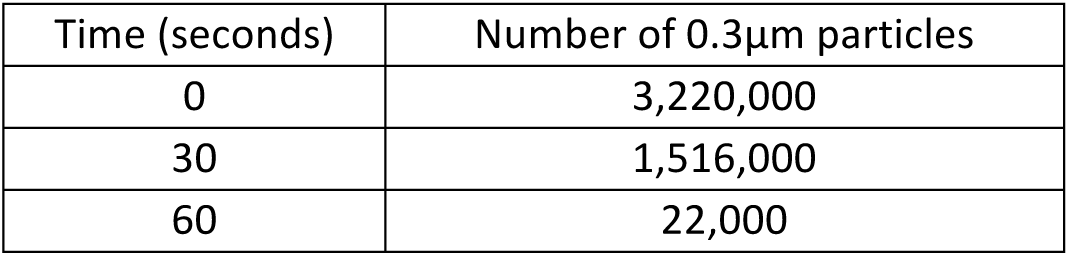
The number of particles detected inside the shield at varying time intervals, with an aspiration unit

## 4 Discussion

### Synopsis of key findings

This study, which assesses the effectiveness of the St George’s shield, is the first to our knowledge to investigate the ability of a physical barrier to reduce aerosol particle spread during a simulated aerosol generating procedure. We found that when using the shield, there was a reduction in the the density of 0.3μm aerosol particles in all theatre staff positions (Table 1a and 1b) and therefore submit that this shield is likely to confer added protection to personnel involved in tracheostomies on patients with COVID-19.

We found that the St George’s shield greatly reduces the number of aerosol particles generated from a simulated aerosol generating procedure, that reach all members of the operating team. (Table 1a and 1b) Our findings suggest that the operating surgeon is exposed to high levels of aerosol particles (36,000 using method 1 and 1,245,000 using method 2) when barrier protection is not used during tracheostomy. The surgeon (location C) was exposed to a notably greater number of aerosol particles when compared to the anaesthetist (an average of 27,500 for methods 1 and 2), scrub nurse (an average of 38,000 for methods 1 and 2) and other theatre personnel (an average of 26,000 for methods 1 and 2). The risk of aerosol particle exposure to the surgeon is greatly mitigated with the use of the shield, to levels comparable with the rest of the theatre environment (a difference of 7000 for method 1 and 1,387,000 for method 2). The difference in aerosol particle detection at location D is also notable (7000 less particles using method 1 and 1,222,000 using method 2). This is important as it represents the location at which the operating surgeon might be leaning over during surgery.

One of our concerns was the release of retained aerosol particles into the immediate theatre environment when the shield is dismantled. Our findings suggest that there is no additional risk of removing the shield following an aerosol generating procedure. Over a period of 10 minutes, the detection of particles (0.3μm, 2.5μm and 10μm) decreased to at least that of the level of the ambient environment. (Table 2) This is important as it illustrates that there is no apparent risk of the particles being contained within the shield and consequently released into the immediate atmosphere once the shield is removed from the patient. Although we estimate that the elapsed time between opening the trachea and finishing the procedure will be greater than 10 minutes, the addition of using an aspiration unit further increases the speed by which particle levels return back to that of the ambient environment (1 minute with an aspiration unit versus 10 minutes without).

Ear, nose and throat surgeons, who perform tracheostomies, are known to be at higher risk of contracting viruses and various COVID-19 guidelines have been published to ensure their safety.^6^ Modifications to standard clinical practice have been paramount to minimise the possibility of exposure to COVID-19 whilst performing these high-risk aerosol generating procedures; such changes include meticulous senior-led discussions about indication, timing and location of tracheostomy, as well as revision of anaesthetic and surgical techniques.^8^ Public Health England guidance emphasises the importance of effective PPE.^10^ In accordance with this, we have shown that the St George’s shield can reduce particle exposure to all theatre staff, especially to the operating surgeon and therefore the St George’s shield has the potential to offer an extra level of PPE to personnel involved in tracheostomies.

### Study limitations

There are limitations to recognise when interpreting the data presented in this study. Firstly, the particle counter detected fixed particle diameters and the smallest particle detected during our experiment was 0.3μm. The aerosol particle size of SARS-CoV-2 has been demonstrated to have a diameter of 0.25μm - 0.5μm.^4, 5^ We are therefore unable to account for viral particles smaller than this size for both our primary and secondary outcome measure; however we are confident that we can extrapolate our data for particles of a similar size to those of SARS-CoV-2. Additionally, the experiment illustrated that over time, the level of particles (0.3μm, 2.5μm and 10μm) decreased to ambient environment levels or below. It is therefore likely that particles smaller than specifically tested during the experiment would follow the same pattern.

A second unknown is how the density of these aerosol particles will vary in different temperatures. We used the particle detector to measure the ward temperature (23.5°C) – this was consistent throughout the experiment. Whilst we do not anticipate that a different temperature in theatre would impact the distribution of aerosol particles, it is possible that the rate of particle dissipation may increase in higher ambient temperatures.

Finally, the accuracy of a cough to mimic the expulsion of SARS-CoV-2 aerosol particles during a tracheostomy is uncertain and may have case-by-case variation depending on a variety of factors, including a patient’s viral load, levels of bronchospasm and concentration of muscle relaxant used.

## 5 Conclusion

We present the St George’s Covid shield, a polycarbonate barrier enclosure, designed as an additional precautionary measure to be used by ENT surgeons performing tracheostomies, to further minimise the transmission of COVID-19 from infected patients to healthcare staff. The clinical simulation study illustrates a significant decrease in the number of particles detected at varying locations when the shield is used, in the presence of a coughing person used to simulate the release of droplets akin to when the trachea is opened during surgery. Results also demonstrate that over time, aerosol particles dissipate, with particle numbers decreasing to ambient levels.

We anticipate that the St George’s shield can be adapted for use during other surgical procedures and in outpatient settings, which will be of vital importance as we continue to operate during the COVID-19 pandemic.

## Data Availability

The authors confirm that the data supporting the findings of this study are available within the article.

